# Validation of an in vitro diagnostic test for endometriosis: impact of confounding medical conditions and lesion location

**DOI:** 10.1101/2024.04.17.24305952

**Authors:** Elza Daoud, David F. Archer, Bárbara Herranz-Blanco

**Affiliations:** Chemo Research, Madrid, Spain; Department of Obstetrics and Gynecology, Eastern Virginia Medical School, Norfolk, Virginia, USA

**Keywords:** In vitro diagnostic test, endometriosis, validation, lesion location, superficial endometriosis, confounding conditions

## Abstract

With the aim to shorten the time for diagnosis and accelerate access to correct management, a non-invasive diagnostic test for endometriosis was developed and validated. The IVD test combines an ELISA test kit to quantify CA125 and BDNF concentrations in serum and a data treatment algorithm hosted in medical software processing results from the ELISA test and responses to six clinical variables. Serum samples and clinical variables extracted from psychometric questionnaires from 77 patients were collected from the Oxford Endometriosis CaRe Centre biobank (UK). Case/control classification was performed based on laparoscopy and histological verification of the excised lesions. Biomarkers serum concentrations and clinical variables were introduced to the software, which generates the qualitative diagnostic result (“positive” or “negative”). This test allowed the detection of 32% of cases with superficial endometriosis, which is an added value given the limited efficacy of existing imaging techniques. Even in the presence of various confounding medical conditions, the test maintained a specificity of 100%, supporting its suitability for use in patients with underlying medical conditions.

## 1. Introduction

Endometriosis is a progressive, estrogen-dependent disease that affects approximately 10% of women of reproductive age [1]. It is characterized by the presence of endometrial-like tissue outside the uterus, commonly affecting the pelvic cavity, ovaries, fallopian tubes, and other surrounding structures[2]. These lesions result in a chronic inflammatory response, which can lead to the formation of scar tissue and adhesions[3]. The clinical presentation of endometriosis can be very diverse, with a wide range of symptoms, including chronic non-menstrual pelvic pain, dysmenorrhea, dysuria, infertility, and many others; with the onset of symptoms usually occurring during adolescence[1,4]. The severity and manifestation of symptoms can be influenced by various factors, including the location and extent of the endometrial implants, hormonal fluctuations, and individual pain thresholds[5,6]. Also, symptoms often overlap with those of various other conditions [7,8]. Although imaging techniques such as transvaginal ultrasound (TVUS) and magnetic resonance imaging (MRI) have been shown to accurately diagnose some endometriosis cases, these are usually limited to more severe stages of the disease[9,10]. Laparoscopy, with or without histological confirmation, remains the gold standard for diagnosing endometriosis, but its invasive nature contributes to diagnostic delays [1–3,11]. Therefore, despite its high prevalence, accurately diagnosing endometriosis can be challenging, with an initial misdiagnosis in up to 65% of women and a diagnostic delay of 4-11 years [7,12].This delay hinders the identification of early stages, allowing the condition to progress, leading to increased severity, fibrosis, and potential infertility [13,14]. Developing a non-invasive diagnostic test for endometriosis becomes crucial in order to obviate the delay in diagnosis [15,16].

Prior studies have delved into an extensive array of biomarkers, highlighting the complexity of understanding endometriosis. CA125, a widely recognized glycoprotein, has been a focal point in research due to its association with various gynecological conditions, including endometriosis [17–19]. Despite its usefulness, the lack of specificity and limited sensitivity as a standalone marker, along with its utility being limited to late stages of the disease, has underscored the need for complementary biomarkers. Brain-derived neurotrophic factor (BDNF), known for its involvement in neuroplasticity and neuronal survival, has emerged as a promising candidate, with studies demonstrating elevated levels in patients with endometriosis compared to healthy controls [11,20,21]. However, the lack of specificity among individual biomarkers emphasizes the necessity of a comprehensive diagnostic approach integrating multiple markers to enhance accuracy and reliability in endometriosis detection.

Recently, we have developed a diagnostic treatment algorithm that combines CA125 and BDNF measurements with six pertinent clinical variables: patient’s surgical history related to endometriosis, the manifestation of painful periods as a leading symptom for endometriosis referral, the intensity of menstrual pain during the previous cycle, the age at the onset of intercourse-related pain, the age at the initiation of regular pain-killer usage, and the age at the initial diagnosis of an ovarian cyst. CA125, BDNF, and the six clinical factors were integrated into the final logistic regression model, achieving an AUC of 0.867, sensitivity of 51.5%, and specificity of 95.6% [22].

The influence of confounding conditions on the final diagnosis of endometriosis using this test was challenged. This is because multiple conditions, gynecological (for instance, adenomyosis [23–26], pelvic inflammatory disease (PID) [27–29], uterine fibroids [29–31] and ovarian cysts [29,32]) and non-gynecological (for instance, inflammatory bowel disease (IBD) [33] or rheumatoid arthritis [34–36], asthma [37], anxiety and depression [38–40]) could affect the levels of CA125 and BDNF.

The primary aim of this study was to validate the diagnostic performance of the test in endometriosis patients while also discerning the specific subgroup of patients in which the test demonstrates superior performance. The secondary aim was to further investigate how confounding conditions influence CA125 and BDNF and whether or not the performance of the test is affected.

## 2. Results

### 2.1. Diagnostic performance by endometriosis lesion type

One hundred percent of controls from the validation dataset were correctly diagnosed (negative) with the IVD test, based on the threshold established in the development dataset. With this, a sensitivity (after weighing for disease stages) of 46.2% (95% CI: 25.5-66.8%) and a specificity of 100% (95% CI: 86.7-100%) was obtained. The accuracy was 64.1% (95% CI: 50.4-77.8%) and the AUC was 0.758 (95% CI: 0.650-0.867). To understand in which subgroup of endometriosis patients the test works best, i.e., is capable of detecting the highest number of cases, patients were separated in subgroups by lesion types. First, the association between the stages of endometriosis and the types of endometriosis lesions was examined using Pearson’s chi-squared test. The analysis revealed a significant association (χ^2^ = 765.76, df = 25, p < 0.001), indicating a strong relationship between the rASRM stages classification of endometriosis and classification by types of lesions. The contingency table (Table 1) provides insight on how lesion types are distributed by endometriosis rASRM stage for patients of pooled development and validation datasets. Superficial lesions are observed mostly in stage I (81.5%). Extended lesions (endometrioma+DIE) are as expected mostly observed in stage IV.

**Table 1.** Contingency table for the distribution of lesion types by endometriosis rASRM stages.

| rASRM stage lesion type | Stage I (n=81) | Stage II (n=30) | Stage III (n=34) | Stage IV (n=41) | Unclassified (n=2) |
| --- | --- | --- | --- | --- | --- |
| C-section | 0 | 0 | 0 | 0 | 2 |
| DIE | 7 | 17 | 7 | 9 | 0 |
| Endometrioma | 5 | 0 | 16 | 7 | 0 |
| Endometrioma + DIE | 0 | 3 | 10 | 20 | 0 |
| Superficial | 66 | 10 | 1 | 2 | 0 |
| Unclassified | 3 | 0 | 0 | 3 | 0 |

Sensitivity was investigated by lesion type. Results (as reported in table 2) indicate that the IVD test successfully identified around half of the cases of DIE and endometrioma. Furthermore, with a sensitivity of 69.70%, the IVD test demonstrate that the test works best in identifying cases of DIE+endometrioma. Interestingly, 32% of cases of superficial endometriosis were correctly identified with the test. As expected, the two cases of endometriosis located within c-section scars could not be identified with the test (different pathophysiology, as described above).

**Table 2.**
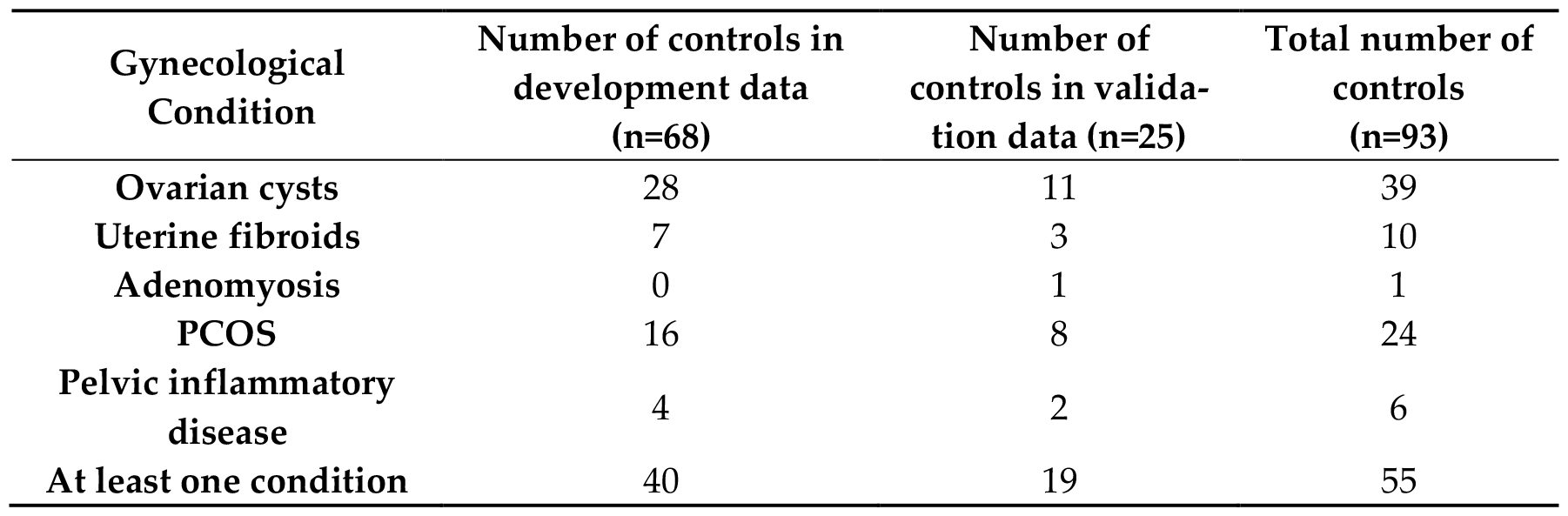
Distribution of cases, number of true positive and sensitivity by lesion type in both development and validation datasets.

An ANOVA was conducted to examine the differences in CA125 values among various types of endometriosis lesions in the pooled datasets (development and validation datasets, figure 1). The results revealed a significant effect of lesion type on CA125 levels (F(5, 275) = 26.162, p < 0.001). Post hoc analyses indicated that the differences were statistically significant (p < 0.001) across the various lesion types. The Tukey multiple comparison of means at a 95% family-wise confidence level revealed several significant differences between the types of lesions in terms of CA125 levels: comparing endometrioma to DIE, there was a statistically significant difference (p<0.01). Additionally, the mean CA125 level (56.05 IU/mL, SD=39.35) were higher for endometrioma than for DIE (32.28 IU/mL, SD=32.69) (p=0.01). Moreover, the mean CA125 level for endometrioma + DIE (67.69 IU/mL, SD=45.49) was higher than the mean CA125 level for DIE (mean=32.28 IU/mL, SD=32.69) (p<0.001). Lower CA125 levels were observed for superficial lesions (mean=19.55 IU/mL, SD=24.74) than for endometrioma (p<0.001), DIE (p=0.02) and endometrioma + DIE (p<0.001). An ANOVA conducted on BDNF values across different lesion types did not show any significant differences of BDNF across different types of lesions (p=0.094). This suggests that the improved sensitivity for DIE+endometrioma lesions is likely to be due to higher levels of CA125 in those lesions, contributing to a higher rate of true positive results in cases with those lesions.

**Figure 1.**
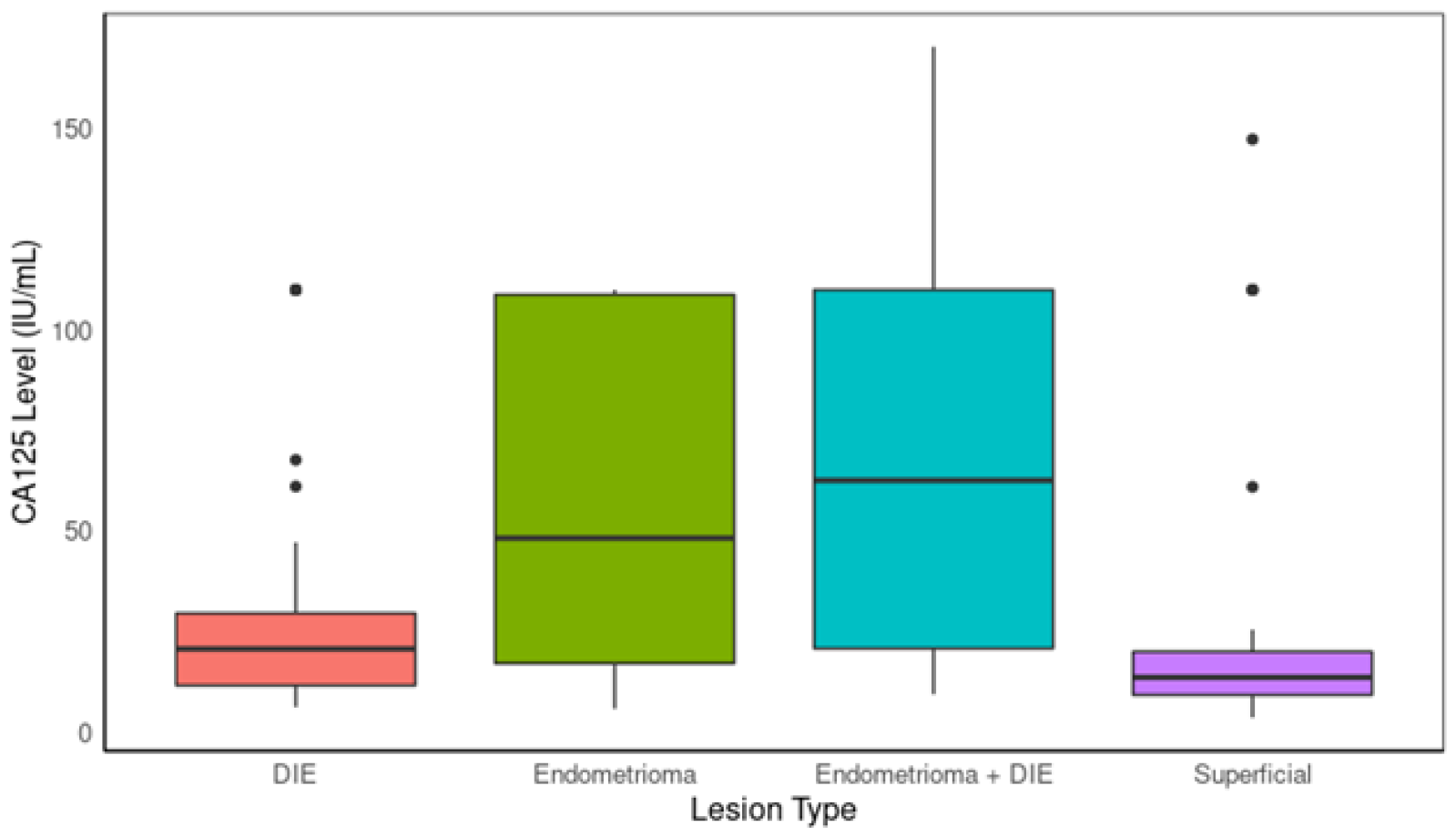
Comparison of CA125 levels between lesion types.

### 2.2. Interference of potentially confounding medical conditions

As shown in table 3, despite 76% (19 out of 25 controls) of controls in the validation dataset having at least one condition that could elevate CA125, the specificity of the diagnostic test was 100%.

**Table 3.**
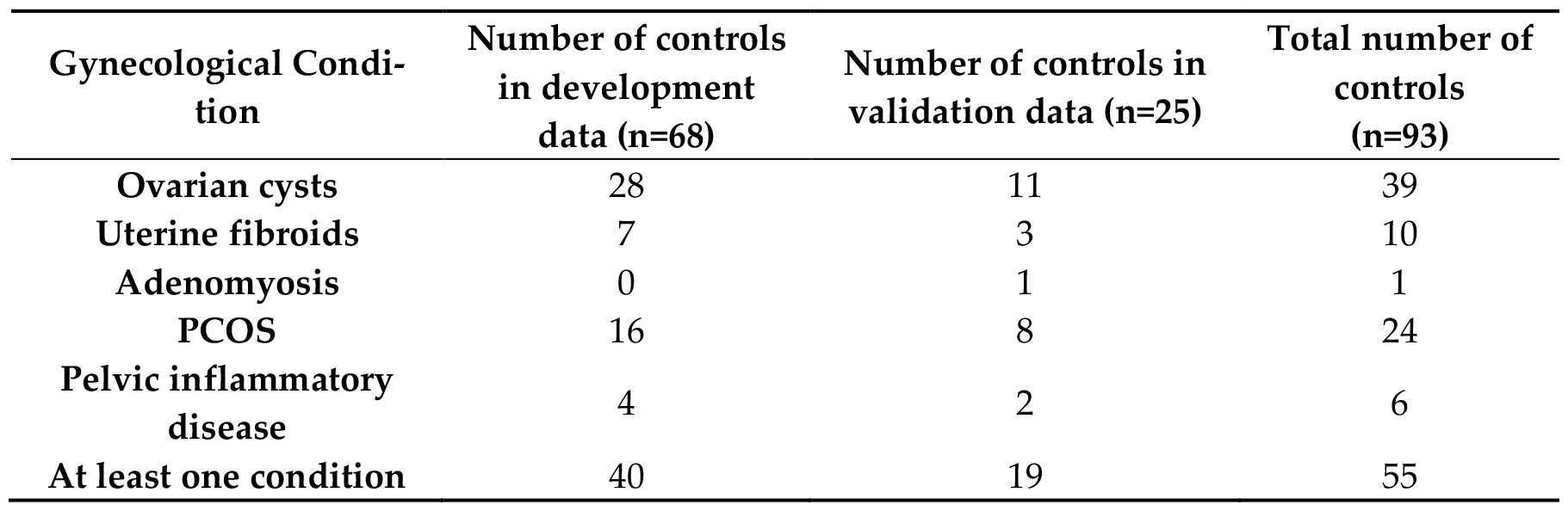
Distribution of gynecological conditions known to elevate CA125 across controls.

Two-way ANOVA with EndoState (Cases/controls) and each confounding condition as predictors was run on CA125 levels in the pooled datasets. For ovarian cysts, the ANOVA revealed the main effect of EndoState (F = 32.97, p<0.001) and Ovarian cyst condition (F = 22.65, p<0.001) on CA125 levels. Individuals with ovarian cysts had higher CA125 values than individuals without ovarian cysts (p<0.001). No interaction between both predictors was reported. For uterine fibroids (UF), a main effect for condition on CA125 was observed (F=11.22, p<0.001) as well as an expected main effect for EndoState (F=15.30, p<0.001). No interaction between both predictors was reported. Individuals with uterine fibroids had higher CA125 values than individuals without uterine fibroids (p<0.001).

Two-way analysis of variance (ANOVA) with EndoState (Cases/controls) and each confounding condition as predictors was run on BDNF in pooled datasets. For Chronic fatigue only, a main effect was observed for EndoState (F=5.75, p=0.017) and an interaction between EndoState and the condition (F=4.20, p=0.04). Pairwise comparisons revealed that cases without chronic fatigue have higher BDNF values than controls with chronic fatigue (mean difference=6.06, p=0.04).

The performance of the diagnostic test was determined in the validation dataset excluding each confounding condition at a time. Results, as shown in Table 4, indicate that the sensitivity values when excluding conditions stay within the 95% CI of the original sensitivity (all conditions included) between 34.3 and 62.9, meaning that no condition critically affects the ability of the test of detecting cases.

**Table 4.**
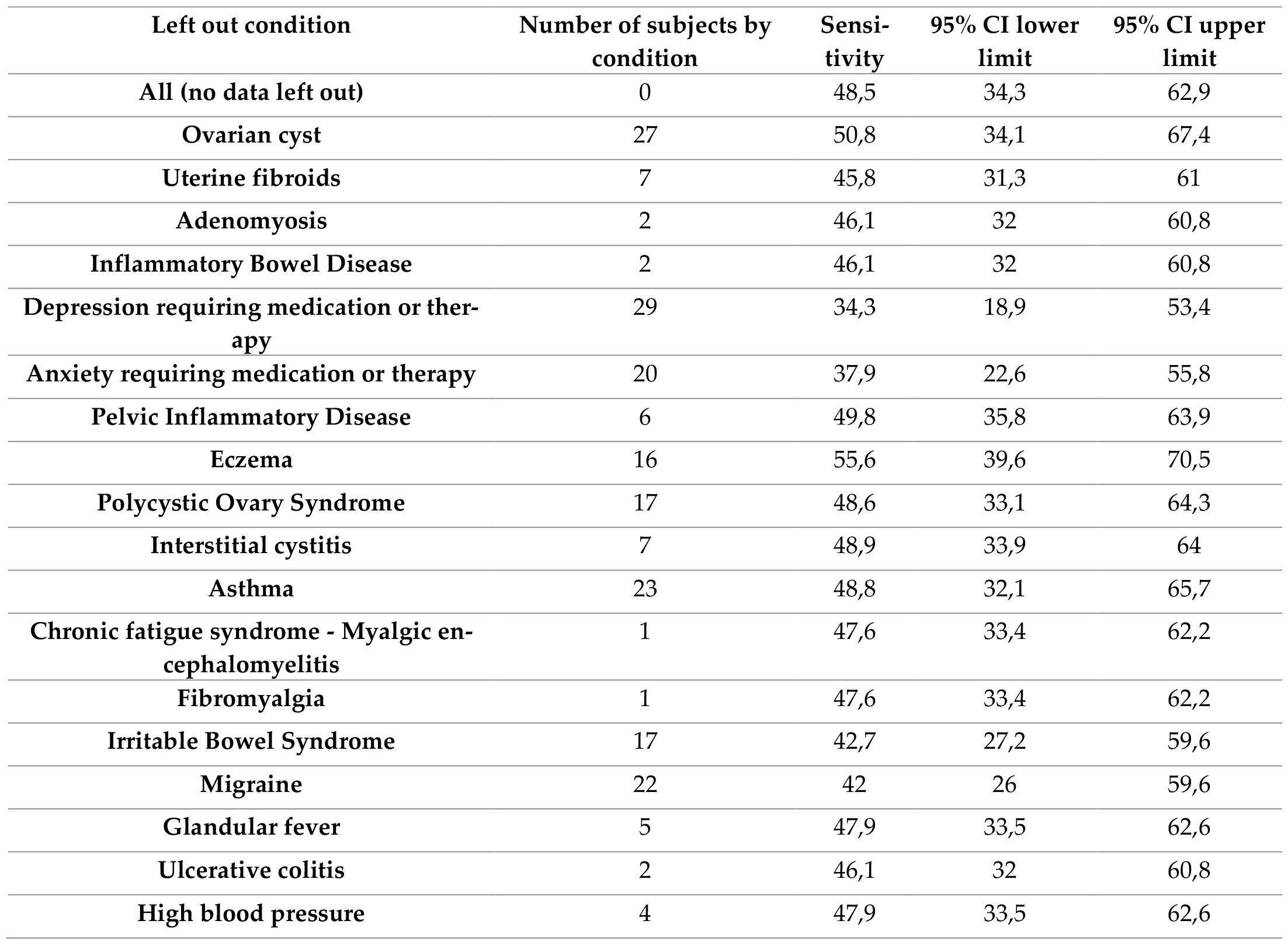
Performance of the IVD test (validation dataset) excluding each medical condition at a time.

## 3. Discussion

The newly developed test for endometriosis demonstrates a high specificity of 100%, suggesting its potential use as a rule-in test in clinical practice. This diagnostic test could significantly contribute to the initial diagnostic workup, effectively confirming the presence of endometriosis and providing clinicians with a reliable tool for early detection and intervention. Moreover, the test demonstrates an encouraging ability to identify superficial lesions of endometriosis, as evidenced by the reported sensitivity of 32 %. This feature is of particular significance considering the constraints associated with the ability of alternative diagnostic methods to detect superficial lesions: superficial endometriosis, characterized by its subtle and less invasive nature, presents unique challenges for detection using ultrasound or MRI. Peritoneal implants invading less than 5 mm of depth from the peritoneal surface are often invisible on MRI [10]. These imaging techniques may struggle to capture the nuanced characteristics of these lesions due to their limited ability to visualize subtle changes in the peritoneum and pelvic surfaces [42]. Additionally, the lack of specific imaging markers or distinguishing features that differentiate superficial lesions from surrounding healthy tissue makes it difficult to accurately identify these lesions using standard imaging modalities. The intricate anatomical location of superficial lesions, often nestled within complex pelvic structures, further contributes to the complexity of their detection, as these areas may be challenging to access and visualize accurately using traditional imaging approaches [43]. By enabling the identification of superficial lesions, the test offers clinicians an essential means of identifying cases that would otherwise have gone undetected, thereby facilitating a more comprehensive and accurate patient management.

Also, the test demonstrated a relatively high sensitivity of 69.70% in detecting endometrioma+ DIE lesions, possibly correlated to patients with those lesions having the highest level of CA125 compared to other types of lesions. Endometrioma, an endometriosis-related ovarian cyst, often exhibits elevated CA125 levels due to its involvement of the ovaries and resulting inflammatory processes. The higher mean CA125 level observed in this group aligns with prior studies [44]. The observed higher mean CA125 level in the endometrioma + DIE lesions compared to the DIE alone, along with the lowest CA125 levels in the superficial endometriosis, suggest that CA125 expression increases with the extent of the disease (i.e., the extent of tissue involvement and disease spread).

Even in the presence of various confounding medical conditions, the test maintains its robustness and reliability, emphasizing its independence from potential confounding factors with 100% of the controls being negative. This characteristic supports its suitability for use in various clinical settings, irrespective of the patient’s medical history, thereby ensuring its applicability without contraindications.

## 4. Materials and Methods

### 4.1. Patients’ characteristics and classification

The current report is a prospective analysis study using biobank samples. A total of 281 samples extracted from the renowned Oxford Endometriosis CaRe Centre biobank in the UK were included for the development (I) and external validation (II) studies. The biobank’s repository comprised meticulously curated serum samples and comprehensive clinical information derived from pre-surgical assessments and post-operative procedures of patients within reproductive age (18–50 years old) undergoing laparoscopy because of a suspicion of endometriosis. Patients were classified as cases or controls based on laparoscopy and thorough evaluation of histological findings. After undergoing laparoscopy, patients diagnosed with endometriosis were categorized into stages according to the revised American Society of Reproductive Medicine (rASRM) classification. Patients who had not used hormones in the 3 months prior to surgery were selected.

136 endometriosis cases and 68 controls were included in the development study (n=204). For the validation study (n=77), 52 cases and 25 controls were included. The demographic characteristics of those patients are available in Table 5. The experimental procedures received approval from the Ethics Committee of CEIm HM Hospitales (codes: 19.05.1411-GHM and 22.03.2001-GHM).

**Table 5.** Demographic characteristics and rASRM classification of the patients in the development (I) and validation (II) studies.

|  | Development study (I) |  | Validation study (II) |  |
| --- | --- | --- | --- | --- |
|  | Controls<br>N=68 | Cases<br>N=136 | Controls<br>N=25 | Cases<br>N=52 |
| **Age years (mean ± SD)** | 33.5 (5.96) | 35.6 (6.42) | 35 (6.44) | 35 (6.47) |
| **BMI (mean ± SD)** | 25.38 (4.63) | 26.46 (5.32) | 26 (5.23) | 26 (5.14) |
| **rASRM classification** |  |  |  |  |
| **I–II** | - | 68 (50%) | - | 42 (81%) |
| **III–IV** | - | 68 (50%) | - | 7 (13%) |
| **Missing information** |  | - | - | 3 (6%) |

### 4.2. Lesion location and subtyping

Imaging findings and surgical examinations have been reported for each subject included in the study. Endometriosis lesions were investigated by location. From these findings, endometriosis lesions were classified into subgroups according to their location in the ovaries and the peritoneal cavity: superficial (< 5 mm depth), endometrioma, and/or deep infiltrative endometriosis (DIE). Specifically, the designation “superficial” was assigned when only superficial endometriosis lesions were identified in the ovaries or peritoneal cavity. The classification of “endometrioma” was used when endometriomas were detected in the ovaries, either with or without accompanying superficial endometriosis. In cases where infiltrative lesions were observed in the peritoneal cavity, with or without associated superficial endometriosis, lesions were classified as “DIE”. Moreover, the “endometrioma + DIE” classification was assigned when both DIE and endometriomas were found in the peritoneal cavity, with or without superficial endometriosis. While endometriosis is thought to be caused by retrograde menstruation, the most likely cause of caesarean section (c-section) scar endometriosis is iatrogenic implantation. Due to this different aetiology, 2 patients with c-section scar endometriosis were misclassified as they should fall under a different category than endometriosis with spontaneous implantation. The distributions of cases of the development and validation studies by lesions type are described in Table 6.

**Table 6.** Classification of endometriosis cases according to lesion location.

| Endometriosis Classification | Development study (I)<br>N=136 | Validation study (II)<br>N=52 |
| --- | --- | --- |
| Superficial | 54 (39.7%) | 24 (46.2%) |
| Endometrioma | 25 (18.4%) | 3 (5.8%) |
| DIE | 28 (20.6%) | 13 (25%) |
| DIE + endometrioma | 25 (18.4%) | 8 (15.4%) |
| Unclassified | 4 (2.9%) | 2 (3.8%) |
| C-section scar | 0 | 2 (3.8%) |

### 4.3. Confounding disease screening

Patients were asked to fill out a presurgical survey including a question to indicate the absence/presence of confounding medical conditions from a list. They were asked: please mark whether you have had any of the following medical conditions, and at what age you were first diagnosed by a doctor (please tick all that apply)” and were given the list of medical conditions. Patients were also asked to indicate whether they were affected by other unlisted medical conditions. This survey was administered to patients in one of its 3 versions: version #1 did not list 3 medical conditions: Anxiety (1), cardiovascular disease (2) and high blood pressure (3). These conditions were only listed in questionnaires #2 and #3. Versions #2 and #3 were responded by 141 out of 190 patients included in the development study (14 patients did not answer to this question out of 204) and 64 out of 77 patients included in the validation study. For completeness, imaging and surgical findings were used to further identify patients with gynaecological conditions.

Table 7 depicts the prevalence of the confounding conditions in patients included in the development and validation studies.

**Table 7.** Prevalence of confounding conditions in the development and validation datasets.

| Confounding condition | Prevalence in development study | Prevalence in validation study |
| --- | --- | --- |
| Anxiety requiring medication or therapy | 39/141 (28%) | 20/64 (31%) |
| Asthma | 42/190 (22%) | 23/77 (30%) |
| Adenomyosis | 7/190 (3.7%) | 2/77 (2.6%) |
| Cardiovascular disease | 0 | 0 |
| Crohn's disease | 0 | 0 |
| Chronic fatigue syndrome - Myalgic encephalomyelitis | 10/190 (5.2%) | 1/77 (1.3%) |
| Depression requiring medication or therapy | 68/190 (35.8%) | 29/77 (38%) |
| Diabetes requiring diet control | 3/190 (1.6%) | 0 |
| Diabetes requiring insulin or tablets | 1/190 (0.5%) | 0 |
| Eczema | 32/190 (16.8%) | 16/77 (21%) |
| Uterine fibroids | 28/190 (9.5%) | 7/77 (9.1%) |
| Fibromyalgia | 4/190 (2%) | 1/77 (1.3%) |
| Glandular fever | 17/190 (8.9%) | 5/77 (6.5%) |
| Graves's disease | 0 | 0 |
| Hashimoto's disease | 0 | 0 |
| High blood pressure | 9/141 (6%) | 4/64 (6.2%) |
| Irritable bowel syndrome | 43/190 (23%) | 17/77 (22%) |
| Interstitial Cystitis | 12/190 (62.5%) | 7/77 (9%) |
| Migraine | 51/190 (27%) | 22/77 (28.6%) |
| Mitral valve prolapse | 2/190 (1%) | 0 |
| Multiple sclerosis | 2/190 (1%) | 0 |
| Ovarian cysts | 93/190 (49%) | 27/77 (35.1%) |
| Pelvic inflammatory disease | 13/190 (6.84%) | 6/77 (7.8%) |
| Polycystic ovarian syndrome | 32/190 (16.8%) | 17 (22.1%) |
| Rheumatoid arthritis | 0 | 0 |
| Sjogren's syndrome | 0 | 1/77 (1.3%) |
| Systemic lupus erythematosus | 0 | 0 |
| Thyroid disease | 3/190 (1.6%) | 0 |
| Ulcerative colitis | 1/190 (0.5%) | 2/77 (2.6%) |

### 4.4. Blood sample collection and biomarkers measurement

The specimens were gathered and managed with explicit patient consent, following the guidelines outlined in the Standard Operating procedures of the World Endometriosis Research Foundation [41]. Before the collection of blood, patients were instructed to maintain a minimum fasting period of 10 hours. The serum samples were then preserved in the biobank at temperatures as low as -80 ºC for a duration of up to 5 years, after which they were transferred to the designated laboratory for analysis. The ELISA utilized in this in vitro diagnostic test functions as a solid-phase sandwich enzyme-immunoassay for the precise determination of BDNF and CA125 levels within human serum [22].

### 4.5. Data treatment algorithm

All the necessary input parameters, including serum CA125, serum BDNF, and clinical variables were gathered. Subsequently, laboratory technicians input this data into the IVD test diagnostic medical software, which houses the data treatment algorithm. The algorithm processed the input and generated outcomes, classifying them as either positive or negative based on whether the value exceeded or fell below the predetermined threshold value, respectively.

### 4.6. Statistical analysis

Statistical analysis was conducted utilizing R software, version 4.1.3, provided by the R Foundation for Statistical Computing in Vienna, Austria. The statistical significance level was set at p < 0.05, indicating a threshold below which results were considered statistically significant. In the validation study, the IVD test software was utilized to compute algorithm scores and their corresponding outcomes. These outcomes were delineated as positive diagnosis when the score surpassed the defined cut-off, and negative diagnosis when the score fell below the defined cut-off. Specifically, the validation study’s sensitivity and specificity were expected to align with or exceed the lower limits of the sensitivity and specificity 95% confidence intervals outlined in the algorithm development study: an AUC of 0.867 with a sensitivity of 51.5% (42.8 - 60.1) at a specificity of 95.6% (86.8 - 98.9%) as reported by Herranz et al. To assess the IVD test clinical performance, the results of the primary performance parameters (sensitivity and specificity) were contrasted with the acceptance criteria values established during the development study. To ensure equitable representation of both the low-stage and high-stage groups, the outcomes in the validation were appropriately weighted.

To further elucidate the performance of the IVD test, the sensitivity for each endometriosis classification, with the values specified alongside their respective 95% CI are reported for the distinct subgroups based on lesion types. For a more comprehensive assessment of test’s efficacy over a larger sample size, development and validation datasets were pooled. Analysis was run on pooled dataset. BDNF values in pooled datasets followed a normal distribution and CA125 values were arithmetically transformed to follow a normal distribution. To investigate the effect of confounding diseases on biomarkers levels and the performance of the test, only conditions with >1% prevalence in both datasets were considered. A two-way ANOVA analysis was conducted to assess the effect of medical conditions and EndoState (Cases/controls) on CA125 and BDNF, respectively, including an interaction term. Only conditions showing significant main effects or interaction will be reported. Furthermore, the performance of the algorithm on validation data was evaluated after excluding each specific conditions, one at a time.

## 5. Conclusions

Overall, the high specificity of the test, coupled with its independence from potential confounding medical conditions, position it as a valuable and reliable tool for the accurate and timely diagnosis of endometriosis.

## 6. Patents

There is a patent resulting from the work reported in this manuscript.

## Data Availability

All data produced in the present study are available upon reasonable request to the authors

## Author Contributions

For research articles with several authors, a short paragraph specifying their individual contributions must be provided. The following statements should be used “Conceptualization, E.D. and B-H.-B; methodology, E.D. and B-H.-B; software, E.D. and B-H.-B; validation, E.D. and B-H.-B.; formal analysis, E.D. and B-H.-B; investigation, E.D. and B-H.-B; resources, E.D and B-H.-B.; data curation, E.D. and B-H.-B.; writing—original draft preparation, E.D.; writing—review and editing, E.D., B.H.-B, D.A.; visualization, E.D..; supervision, B.H.; project administration, E.D. and B-H.-B.; funding acquisition, E.D. and B-H.-B. All authors have read and agreed to the published version of the manuscript.” Please turn to the [CRediT taxonomy](https://img.mdpi.org/data/contributor-role-instruction.pdf) for the term explanation. Authorship must be limited to those who have contributed substantially to the work reported.

## Institutional Review Board Statement

The study was conducted in accordance with the Declaration of Helsinki, and approved by the Institutional Review Board (or Ethics Committee) of CEIm HM Hospitales (codes: 19.05.1411-GHM and 22.03.2001-GHM and date of approval: April 12^th^, 2022

## Informed Consent Statement

Informed consent was obtained from all subjects involved in the study. The experimental procedures received approval from the Ethics Committee of CEIm HM Hospitales (codes: 19.05.1411-GHM and 22.03.2001-GHM).

